# Reduced Incidence of Long-COVID Symptoms Related to Administration of COVID-19 Vaccines Both Before COVID-19 Diagnosis and Up to 12 Weeks After

**DOI:** 10.1101/2021.11.17.21263608

**Authors:** Michael A. Simon, Ryan D. Luginbuhl, Richard Parker

## Abstract

Both clinical trials and studies leveraging real-world data have repeatedly confirmed the three COVID-19 vaccines authorized for use by the Food and Drug Administration are safe and effective at preventing infection, hospitalization, and death due to COVID-19 and a recent observational study of self-reported symptoms provides support that vaccination may also reduce the probability of developing long-COVID. As part of a federated research study with the COVID-19 Patient Recovery Alliance, Arcadia.io performed a retrospective analysis of the medical history of 240,648 COVID-19-infected persons to identity factors influencing the development and progression of long-COVID. This analysis revealed that patients who received at least one dose of any of the three COVID vaccines prior to their diagnosis with COVID-19 were 7-10 times less likely to report two or more long-COVID symptoms compared to unvaccinated patients. Furthermore, unvaccinated patients who received their first COVID-19 vaccination within four weeks of SARS-CoV-2 infection were 4-6 times less likely to report multiple long-COVID symptoms, and those who received their first dose 4-8 weeks after diagnosis were 3 times less likely to report multiple long-COVID symptoms compared to those who remained unvaccinated. This relationship supports the hypothesis that COVID-19 vaccination is protective against long-COVID and that effect persists even if vaccination occurs up to 12 weeks after COVID-19 diagnosis. A critical objective of this study was hypothesis generation, and the authors intend to perform further studies to substantiate the findings and encourage other researchers to as well.

## Background

Clinical trials and studies of real-world evidence have demonstrated repeatedly that the three COVID-19 vaccines* authorized for emergency use or approved for use by the Food and Drug Administration are safe and effective at preventing COVID-19 infection, hospitalization, and death (*1*). A recent observational study of self-reported symptoms goes further, concluding that vaccination may also reduce the likelihood and intensity of long-COVID (*2*), a proposal that has been raised anecdotally, as well (*3*). The COVID-19 Patient Recovery Alliance is performing a federated real-world data study of long-COVID and as a part of the federated study Arcadia.io performed a retrospective analysis of the medical history of COVID-19-infected persons to identify factors, both before and after COVID-19 diagnosis, influencing the development and progression of long-COVID.

## Methods

Data for this analysis were collected from Arcadia Data Research (Arcadia.io, Burlington, MA), a normalized, de-identified clinical and operational dataset containing over 150 million patient records. Data were captured directly from electronic health record (EHR) systems, practice management systems, and health care payer claims and eligibility data, and subjected to data quality analyses for compliance with quality measure, risk adjustment, utilization and finance, and care management requirements. The MITRE Institutional Review Board (IRB) reviewed the study submission, COVID-19 Analytics (MIRB 2020017), and found that it is exempt under the provisions of 45 CFR 46.104(d)(4) as secondary research and approved the study. For the purposes of this analysis, all patients needed to meet the following criteria: (1) patient must have been alive as of March 1, 2020; (2) patient must have had at least one encounter with a provider documenting patient history prior to January 1, 2020; and (3) patient must have had at least one encounter with a practitioner who evaluated their health status after January 1, 2020. All qualifications needed to have been met by May 31, 2021, the cutoff for the data extract. This yielded a data sample covering roughly a year’s worth of COVID-19 activity in the United States, about six months of vaccination activity, and a consistent snapshot of the pandemic prior to the emergence of the delta variant (B 1.617.2) as the predominant variant in the United States at the end of June 2021 (*4*).

Patient demographics and health status were determined based on self-reported sex, age, race, and ethnicity, payer-reported eligibility, and clinically-reported medical conditions and symptoms, identified by ICD-9 or ICD-10 codes. Patients qualified for inclusion were further classified as having been diagnosed with COVID-19 during this period or not. The study cohort was further limited to patients diagnosed with COVID-19, having met either of the following requirements: (1) they were diagnosed with ICD-10 code U07.1 at any time or B97.29 prior to May 2020 in a medical encounter (i.e., the diagnosis was assessed by a provider in a face-to-face or equivalent encounter); or (2) they received a positive result from a COVID-19 nucleic acid amplification test (NAAT) or antigen test result. For patients meeting these criteria, an “index date” was set for the first incidence of COVID-19 diagnosis or positive test result. The index date needed to be at least 20 weeks prior to the cutoff date of the data extraction for the patient to be included in the sample population. Patients who died within twelve weeks of this index date were also excluded from this analysis, as any long-COVID outcomes could not be determined for those individuals.

To simplify the analysis, the set of conditions considered here were defined by two ICD-10 value sets: COVID-19 associated conditions (such as diabetes or chronic kidney disease)^§^ and COVID-19 associated symptoms (such as fever or loss of taste and smell)^†^. The value sets were established by the COVID-19 Interoperability Alliance and were provided courtesy of MITRE and Clinical Architecture (*5*). Long-COVID cases were classified as those where the patient presented one or more COVID-associated symptoms between 12 and 20 weeks after the initial COVID-19 diagnosis (*6*). Vaccination data were sourced from payer reports and documentation in EHR systems; no distinction was made between the three U.S. vaccine manufacturers, and although dose sequences were tracked, only the timing of the first dose was used for this analysis.

A logistic regression model based on a Newton Conjugate Gradient solution (Python statsmodels v0.12.2) was used to identify factors potentially influencing the persistence or onset of long-COVID symptoms. The model was applied to each of the long-COVID symptom variables, which included individual symptoms^†^ and two aggregate indicators, “Any Symptom” and “>1 Symptom”. The vaccination timing bins – first dose prior to COVID diagnosis, between 0 and 4 weeks post-diagnosis, between 4 and 8 weeks post-diagnosis, between 8 and 12 weeks post-diagnosis, and unvaccinated prior to 12 weeks post-diagnosis – were used as “one-hot” inputs to the model, leaving the last category (“unvaccinated”) as reference. Patient age and payer type were found to be highly colinear and so were combined into a single factor for statistical analysis^¶^. Patient sex, age and payer combination, race, ethnicity, hospitalization status during acute infection, and pre-existing COVID-associated conditions^§^ were all included as additional factors. The resulting parameters from the model fit were filtered by p-value to α_c_<0.05 and converted to odds ratios to determine effect on risk of exhibiting long-COVID symptoms.

To further test specific outcomes identified using the logistic regression model, a general linear model using an iteratively reweighted least squares optimization method (Python statsmodels v0.12.2) was fitted to an aggregate continuous variable counting the number of distinct long-COVID symptoms reported after 12-weeks following diagnosis (“Symptom Count”). In contrast to the binned vaccination timing variables used with the logistic regression model, vaccination timing in the linear model was defined as a ratio representing the timing of the first vaccination dose relative to infection, where 1=vaccination prior to COVID-19 infection, 0=unvaccinated at 20 weeks post-COVID-19 infection, and the values in between representing 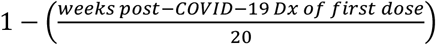. As before, patient sex, age and payer combination, race, ethnicity, and pre-existing COVID-associated conditions were included as inputs to the model, and the resulting coefficient parameters were filtered by p-value to α_c_<0.05.

## Results

Given the initial population inclusion criteria, 25,804,278 persons in the Arcadia Data Research dataset were considered eligible for this study. Between February 2020 and May 2021, 1,065,626 of those persons were diagnosed with COVID-19. Of those patients, 240,648 met the additional inclusion criteria of qualifying provider encounters both before January 1, 2020, and at least 20 weeks after their first COVID-19 diagnosis. Of these patients, 220,460 (91.6%) had not received any vaccine against COVID-19 prior to 12-weeks after their COVID-19 diagnosis, 2,392 (1.0%) received their first dose prior to diagnosis, and the remainder, 17,796 (7.4%), were vaccinated within the first twelve weeks after COVID-19 diagnosis. Vaccination during that 12-week period after COVID-19 diagnosis was further broken down into four-week bins for statistical analysis (Table 1).

**TABLE 1.**
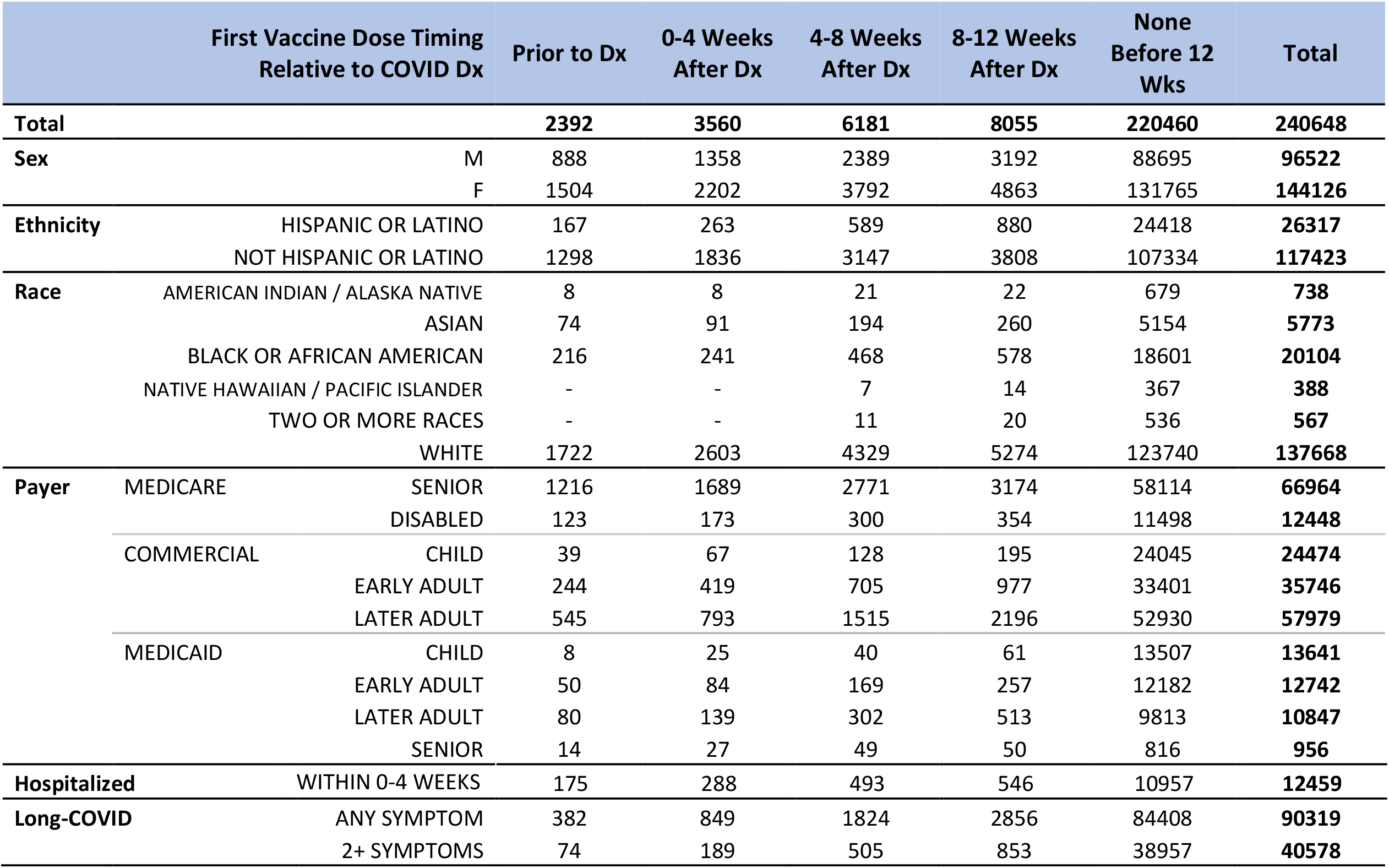
Cohort Demographics – Patient counts by timing of first vaccine dose, categorized by demographic factors. Unknown or Unreported counts are not shown and some small groups or cells were redacted to preserve privacy so values shown may not sum to total patient counts.

Based on the results of the logistic regression model, even a single dose of any of the three COVID-19 vaccines had a protective (<1.0) odds ratio relative to a patient who had no record of vaccination if vaccines were administered up to 12 weeks after diagnosis, and this protective effect was stronger the earlier the first dose occurred relative to COVID-19 infection (Table 2). This result was supported by the general linear model, which also characterized vaccination as having a strongly negative effect on likelihood and number of long-COVID symptoms, representing the strongest parameter in the model (param = -0.85, p-value<0.0005) (Table 3).

**TABLE 2.** Logistic Regression / Odds Ratios – Results of logistic regression models on patients diagnosed with COVID-19 who were vaccinated before COVID-19 infection, within 12 weeks after COVID-19 infection, or remained unvaccinated 12 weeks after COVID-19 infection. “Any Symptom” model estimates the likelihood that the patient will experience any long-COVID symptom, and the “>1 Symptoms” model estimates the likelihood that the patient will experience more than one distinct long-COVID symptom. Reference group for odds ratios is patients who remained unvaccinated after 12 weeks following their first COVID-19 diagnosis. Other factors (not shown) included: Demographic characteristics based on self-reported information, payer information based on reports by health plan payer as of date of infection, indication of whether patient was hospitalized for COVID-19 (determined by diagnosis code) within the first four weeks of diagnosis, and documentation of a COVID-associated chronic conditions prior to date of COVID-19 diagnosis. Vaccination was the most consistent and substantial contributor to the model.

| FIRST VACCINE DOSE TIMING | ANY SYMPTOM |  |  |  | >1 SYMPTOM |  |  |  |
| --- | --- | --- | --- | --- | --- | --- | --- | --- |
|  | *Odds Ratio* | *Likelihood* | *95% CI* | *p-value* | *Odds Ratio* | *Likelihood* | *95% CI* | *p-value* |
| **Prior to COVID-19 diagnosis** | 0.220 | 4.5X | (0.196) – (0.245) | <0.005 | 0.113 | 8.8X | (0.090) – (0.143) | <0.005 |
| **0-4 weeks after diagnosis** | 0.382 | 2.6X | (0.353) – (0.413) | <0.005 | 0.189 | 5.3X | (0.163) – (0.220) | <0.005 |
| **4-8 weeks after diagnosis** | 0.535 | 1.9X | (0.506) – (0.567) | <0.005 | 0.317 | 3.2X | (0.289) – (0.348) | <0.005 |
| **8-12 weeks after diagnosis** | 0.747 | 1.3X | (0.713) – (0.784) | <0.005 | 0.458 | 2.2X | (0.426) – (0.493) | <0.005 |

**TABLE 3.** General Linear Model Results – Results of linear regression model on patients diagnosed with COVID-19 who were vaccinated before COVID-19 infection, within 20 weeks after COVID-19 infection, or remained unvaccinated 20 weeks after COVID-19 infection. Model estimates the number of distinct long-COVID symptoms the infected patient is likely to demonstrate. The Vaccination input is a continuous variable ranging from 0-1, demographic characteristics were based on self-reported information, payer was as of date of infection, hospitalization indicates whether patient was hospitalized for COVID-19 (as indicated by diagnosis code) within the first four weeks of diagnosis, and chronic conditions represent documentation of a COVID-associated chronic condition prior to date of COVID-19 diagnosis. Vaccination was by far the most significant contributor to the model.

| *Category* | *Feature* | *Coef* | *p-value* | *95% CI* |
| --- | --- | --- | --- | --- |
| **Sex** | M | 0.35768 | 0.000 | ( 0.312) - ( 0.402) |
|  | F | 0.52935 | 0.000 | ( 0.484) - ( 0.574) |
| **Ethnicity** | HISPANIC OR LATINO | 0.07936 | 0.000 | ( 0.058) - ( 0.100) |
|  | NOT HISPANIC OR LATINO | 0.00256 | 0.711 | (-0.010) - ( 0.016) |
| **Race** | AMERICAN INDIAN OR ALASKA NATIVE | 0.01928 | 0.713 | (-0.083) - ( 0.122) |
|  | ASIAN | -0.06517 | 0.001 | (-0.103) - (-0.026) |
|  | BLACK OR AFRICAN AMERICAN | 0.09141 | 0.000 | ( 0.067) - ( 0.114) |
|  | NATIVE HAWAIIAN OR OTHER PACIFIC ISLANDER | -0.12427 | 0.083 | (-0.264) - ( 0.016) |
|  | TWO OR MORE RACES | -0.12193 | 0.041 | (-0.238) - (-0.004) |
|  | WHITE | 0.02573 | 0.000 | ( 0.011) - ( 0.039) |
| **Payer** | MEDICARE |  |  |  |
|  | SENIOR | 0.06966 | 0.001 | ( 0.028) - ( 0.111) |
|  | DISABLED | 0.33304 | 0.000 | ( 0.285) - ( 0.380) |
|  | COMMERCIAL |  |  |  |
|  | CHILD | -0.12243 | 0.000 | (-0.167) - (-0.077) |
|  | EARLY ADULT | 0.05719 | 0.010 | ( 0.013) - ( 0.100) |
|  | LATER ADULT | 0.04703 | 0.027 | ( 0.005) - ( 0.088) |
|  | MEDICAID |  |  |  |
|  | CHILD | -0.11636 | 0.000 | (-0.164) - (-0.067) |
|  | EARLY ADULT | 0.19178 | 0.000 | ( 0.143) - ( 0.239) |
|  | LATER ADULT | 0.24079 | 0.000 | ( 0.192) - ( 0.289) |
|  | SENIOR | 0.19211 | 0.000 | ( 0.093) - ( 0.290) |
| **Vaccination** | FIRST DOSE 0-20 WEEKS | -0.85137 | 0.000 | (-0.880) - (-0.821) |
| **Hospitalized** | WITHIN 0-4 WEEKS | 0.06327 | 0.000 | ( 0.036) - ( 0.089) |
| **Conditions** | NO PREVIOUS CONDITIONS | -0.05084 | 0.000 | (-0.070) - (-0.030) |
|  | DIABETES MELLITUS | 0.04582 | 0.000 | ( 0.030) - ( 0.061) |
|  | METABOLIC SYNDROME | 0.03766 | 0.074 | (-0.003) - ( 0.078) |
|  | OBESITY | 0.01366 | 0.066 | (-0.000) - ( 0.028) |
|  | COGNITIVE DISORDERS | 0.28994 | 0.000 | ( 0.262) - ( 0.317) |
|  | CEREBROVASCULAR ACCIDENT | 0.16007 | 0.000 | ( 0.129) - ( 0.190) |
|  | TRANSIENT ISCHEMIC ATTACK | 0.11564 | 0.000 | ( 0.081) - ( 0.149) |
|  | COAGULOPATHIES | 0.18982 | 0.000 | ( 0.164) - ( 0.214) |
|  | CARDIAC ARREST | 0.18890 | 0.000 | ( 0.084) - ( 0.292) |
|  | CARDIAC ARRHYTHMIAS | 0.18267 | 0.000 | ( 0.165) - ( 0.199) |
|  | CARDIOMYOPATHY | -0.04337 | 0.011 | (-0.076) - (-0.009) |
|  | MYOCARDIAL INFARCTION | 0.12337 | 0.000 | ( 0.095) - ( 0.151) |
|  | HEART FAILURE | 0.14064 | 0.000 | ( 0.114) - ( 0.166) |
|  | HYPERTENSION | 0.07047 | 0.000 | ( 0.054) - ( 0.086) |
|  | MYOCARDITIS | 0.25440 | 0.026 | ( 0.030) - ( 0.478) |
|  | VALVULAR HEART DISEASES | 0.09274 | 0.000 | ( 0.072) - ( 0.112) |
|  | NEPHROTIC SYNDROME | 0.28041 | 0.000 | ( 0.139) - ( 0.421) |
|  | GLOMERULONEPHRITIS | 0.06662 | 0.543 | (-0.148) - ( 0.281) |
|  | RENAL FAILURE / CKD | 0.05294 | 0.000 | ( 0.032) - ( 0.073) |
|  | DEPENDENCE ON RENAL DIALYSIS | 0.37229 | 0.000 | ( 0.305) - ( 0.438) |
|  | ASTHMA | 0.19121 | 0.000 | ( 0.175) - ( 0.206) |
|  | CHRONIC OBSTRUCTIVE PULMONARY DISEASE | 0.23238 | 0.000 | ( 0.210) - ( 0.253) |
|  | DEPENDENT ON SUPPLEMENTAL OXYGEN | 0.34735 | 0.000 | ( 0.302) - ( 0.392) |
|  | PULMONARY HYPERTENSION | 0.04825 | 0.010 | ( 0.011) - ( 0.084) |

## Discussion

In this study, patients who had been vaccinated prior to COVID-19 infection were significantly less likely to have long-COVID symptoms. This result applies even if only a single dose of the vaccine is documented, regardless of the manufacturer of the vaccine. Although these results show that other factors, such as demographic factors and chronic conditions, also influence the likelihood that an individual will exhibit long-COVID symptoms, vaccination status had a consistently and substantially larger effect on this outcome than any other factor measured.

Furthermore, patients whose first vaccination occurred within 12 weeks after COVID-19 diagnosis were significantly less likely to have long-COVID symptoms than if they had remained unvaccinated. This finding is consistent with the hypothesis that a vaccine may accelerate clearance of the remaining SARS-CoV-2 virus from specific body compartments or reduce part of the body’s immune response related to development of long-COVID (*3*).

Currently, CDC recommends “vaccination of people with known current SARS-CoV-2 infection should be deferred until the person has recovered from the acute illness (if the person had symptoms) and they have met [criteria](https://www.cdc.gov/coronavirus/2019-ncov/hcp/duration-isolation.html) to discontinue isolation,” but there is no specific recommendation of earlier vaccine administration, nor any mention of potential preventative effects on the development of long-COVID (*7*). These results suggest added benefit to post-infection vaccination, and further indicate that the earlier vaccine is administered, the more significant the benefit. If these results are substantiated with further studies, significant public health benefits could be realized, since up to 30% of those infected with SARS-CoV-2 developed prolonged symptoms, with many of these being debilitating and lasting for months.

This study does not address whether COVID-19 vaccines administered within 12 weeks after COVID-19 infection are effective at preventing subsequent COVID-19 infection. Additionally, this study cannot conclude whether COVID-19 vaccines are safe for use for patients in the acute stage of SARS-CoV-2 infection, although no deleterious effects were detected in vaccinated patients in this study or a separate observational study on patients presenting long-COVID symptoms (*8*). However, the reduced likelihood of long-COVID symptoms observed in this study provides a rationale for vaccination sooner rather than later, achieving improved patient health outcomes related to long-COVID.

This study is subject to at least six limitations: First, these findings are based on opportunistic availability of large volumes of patient data and may have geographic, temporal, contractual, and socioeconomic gaps that could influence outcomes. Second, these findings use vaccination data recorded by payer entities or documented in EHRs by providers but do not incorporate dedicated vaccination surveillance data sources and so may have gaps in vaccination data that are presently undetectable. Third, it is possible, but unlikely, that some of the patients with COVID-19 were misclassified due to a false-positive test result (with no documented correction) or an inaccurate COVID-19 diagnosis. Fourth, no distinction was made between which of the three U.S. COVID-19 vaccines administered; it is possible that some of the effect described is related to a specific vaccine, and that such an effect could not be detected based on the data used here. Fifth, while interactions between the observed demographic factors have been explored, interactions between pre-existing chronic conditions have not and may introduce unforeseen effects to the findings described here. And sixth, this analysis was conducted on patient data collected prior to the emergence of the delta variant as the predominant variant circulating in the United States; as a result, we cannot conclude that the protective effect of vaccination against long-COVID described here applies to patients infected with the delta variant. Furthermore, variant specific analysis was not possible as variant data was not available in this dataset.

A critical goal of this study was hypothesis generation, and we intend to perform further studies to substantiate the findings and hope others will as well. Despite the above limitations, the findings reported here support the hypothesis that COVID-19 vaccination reduces the likelihood of developing long-COVID symptoms and does so even if the first dose is received up to 12 weeks post diagnosis. The Long-COVID Study, developed in collaboration with the COVID-19 Patient Recovery Alliance (PRA) and of which this analysis is a part, is intended to offer useful context for COVID-19 vaccine policy development as well as preliminary data for further exploration. To this end, this team and others associated with the PRA will be expanding on this effort to better clarify these results and determine the extent to which these conclusions apply to other circumstances – for example, those around multiple infections and the emergence of the delta variant – efforts that we hope will be joined by others seeking to better understand the causes underlying long-COVID.

## Data Availability

De-identified data used in this study are not available for public viewing.

## Acknowledgements

Mary Kuchenbrod, Jacob Hochberg, Bill Salvucci, Arcadia.io; Dr. Brett Giroir, Data and Evidence Workgroup, Participants in the COVID-19 Patient Recovery Alliance, https://covid19patientrecovery.org/; Anonymous reviewers at MITRE Corporation.

## Footnotes

* The three vaccines approved for emergency use authorization during the study period were the two mRNA vaccines (Pfizer-BioNTech and Moderna) and the inactivated viral vaccine (Johnson & Johnson). No distinction is made in this study between the vaccines and all results only focus on timing of the first dose.

† COVID-19 associated symptoms categorized by body system (value sets supplied by Clinical Architecture as of April 18, 2021): Cardiovascular – Chest Pain, palpitations; Constitutional – Altered mental state, anorexia, chills, fatigue, fever, malaise; Ears, Nose, Mouth, Throat – Loss of sense of smell, loss of sense of taste, nasal congestion, sore throat; Gastrointestinal – Abdominal pain, diarrhea, digestive changes, nausea and/or vomiting; Musculoskeletal – Arthralgia, muscle weakness, general weakness, myalgia; Neurological – Altered mental state, headache; Respiratory – cough, dyspnea.

§ COVID-19 associated conditions categorized by body system (value sets supplied by Clinical Architecture as of April 18, 2021): Cardiac arrest, cardiac arrhythmias, cardiomyopathy, heart failure, history of deep vein thrombosis, hypertension, myocardial infarction, myocarditis, pulmonary hypertension, valvular heart diseases, diabetes mellitus, metabolic syndrome, chronic kidney disease, glomerulonephritis, nephrotic syndrome, dependence on renal dialysis, renal failure, coagulopathies, stroke, cognitive disorders, transient ischemic attack, asthma, chronic obstructive pulmonary disease, dependent on supplemental oxygen, nonspecific respiratory viral infections, obesity.

¶ Age Group/Payer Type combinations used in this analysis were: Medicare – Senior (65+yo), disabled (<60 yo); Commercial – Child (0-24yo), early adult (25-44yo), later adult (45-64yo); Medicaid – Child (0-24yo), early adult (25-44yo), later adult (45-64yo), senior (65+yo).

